# Augmenting Electronic Health Records for Adverse Event Detection

**DOI:** 10.64898/2026.02.10.26345962

**Authors:** Gün Kaynar, Zhaoyi You, Richard D. Boyce, Takahiro Yakoh, Carl Kingsford

## Abstract

**Objective:** Adverse events (AEs) resulting from medical interventions are significant contributors to patient morbidity, mortality, and healthcare costs. Prediction of these events using electronic health records (EHRs) can facilitate timely clinical interventions. However, effective prediction remains challenging due to severe class imbalance, missing labels, and the complexity of EHR records. Classical machine learning approaches frequently underperform due to insufficient representation of minority adverse event classes and limited capacity to capture interactions among patient demographics, administered medications, and associated complications.

**Methods:** We introduce TASER-AE, a novel data augmentation pipeline tailored for structured EHR data, coupled with transformer-based classification. TASER-AE addresses these issues through an NLP-inspired data augmentation framework adapted for EHR, enabling effective minority-class representation in sparse and imbalanced clinical datasets. The augmented records produced by TASER-AE alleviate class imbalance by enriching the representation of minority adverse event classes, which enhances the robustness and predictive performance of the classifier.

**Results:** TASER-AE yields minority-class F1 scores up to 0.70, substantially surpassing classical machine-learning baselines and prior augmentation methods across multiple adverse event tasks. Experiments conducted on two distinct EHR datasets confirm TASER-AE’s ability to substantially improve adverse event detection performance.

**Conclusion:** These results demonstrate the potential of structured, NLP-inspired augmentation methods to overcome data limitations in clinical predictive modeling, ultimately contributing to improved patient safety outcomes. TASER-AE is available at https://github.com/Kingsford-Group/taserae.

## 1 Introduction

Adverse events (AEs) are unintended injuries or complications resulting directly from medical care rather than a patient’s underlying health conditions [33]. AEs may result in prolonged hospitalization, permanent harm, or even death. The prevalence and severity of AEs underscore their critical impact: approximately 25% of hospitalized Medicare patients experienced adverse events during hospitalization [43], with adverse drug events alone responsible for nearly 106,000 deaths annually in the United States [25, 26]. Globally, unsafe medical practices contribute significantly to millions of deaths each year, highlighting the urgent need for effective preventive measures to enhance patient safety and minimize healthcare costs [14, 29, 46]. Predicting AEs in advance offers the potential for timely clinical interventions, significantly improving patient outcomes [35]. Recent advances in machine learning using electronic health record (EHR) data have demonstrated a strong potential for identifying patients who are at a heightened risk for various adverse outcomes, such as acute kidney injury, hospital readmission, and cardiac incidents [1, 24, 28, 31, 40]. Examples include deep learning models that analyze EHR-derived time-series data and have been effectively deployed to estimate continuous risks of clinical deterioration, enabling clinicians to proactively intervene through targeted preventive strategies, such as medication adjustments or intensified patient monitoring [41]. Consequently, enhancing AE prediction models is essential for improving the overall quality of patient care and safety.

EHRs are comprehensive digital repositories of patient medical information, encompassing structured data such as demographics, diagnoses, medications, laboratory results, and unstructured data, including clinical narratives and physician notes [3]. Ideally, these rich datasets should facilitate robust predictive analytics by encapsulating critical risk factors and clinical observations indicative of future adverse events. However, EHR data pose significant analytical challenges due to their inherent heterogeneity, frequent data gaps, inconsistent coding practices across healthcare institutions, and critical clinical information often hidden within unstructured text. These challenges complicate efforts to reliably extract predictive signals, affecting the performance of predictive models [47].

AE prediction is particularly problematic due to fundamental data limitations. Foremost, AEs are relatively infrequent events within healthcare records, leading to a significant class imbalance that hampers machine learning model performance. Furthermore, AE EHR phenotypes are often imprecise and incomplete, with considerable under-reporting and coding inconsistencies complicating accurate ground truth determination [15, 18, 38]. Collectively, these factors result in poor recall (sensitivity), low precision (positive predictive value), and a tendency for model overfitting, diminishing the predictive utility of current AE prediction models [20, 47].

To mitigate similar limitations in other settings, recent research within natural language processing (NLP) has explored data augmentation techniques. Such methods artificially expand training data by creating modified but semantically coherent variants of existing records through straightforward operations such as synonym replacement, random insertion, deletion, and word swapping. By introducing controlled variability, data augmentation broadens the model’s understanding of feature spaces and improves its ability to generalize and accurately predict rare outcomes [34, 44].

In the healthcare domain, data augmentation methods have predominantly been applied to clinical imaging data [9, 16, 37], though recent work has begun extending these techniques to structured EHR data [5]. Ishikawa et al. [21], for instance, introduced an augmentation approach where structured patient data, treated similarly to textual sentences, underwent semantic replacements determined by cosine similarity metrics derived from word embeddings. This approach demonstrated significant improvements, enhancing accuracy and the F1-score notably on highly imbalanced datasets. Likewise, Chen & Du [8] employed self-attentive adversarial augmentation networks (SAAN) to synthesize high-quality minority-class clinical notes, achieving state-of-the-art performance in clinical text classification tasks. These findings affirm that intelligently designed synthetic augmentation methods surpass conventional oversampling techniques in addressing class imbalance and improving AE prediction from structured EHR data [17, 50].

Existing AE prediction baselines range from simple heuristic trigger tools to more sophisticated machine learning classifiers, often limited by low sensitivity and accuracy, particularly for rare-event classes [13, 19]. Without effective data augmentation, typical approaches such as cost-sensitive learning or naive oversampling offer only incremental improvements [45, 49]. Consequently, the exploration of specialized text-based augmentation strategies remains an open area for significant advancement.

We make three main contributions. First, we formulate a broad set of NLP-style token edits for structured, tabular EHR data by operating on tokenized patient summaries and enforcing type-consistent constraints so that edits preserve clinical meaning. Second, we identify a simple and effective augmentation policy for rare adverse events that combines synonym insertion, random token deletion, and shuffling, and we show that this combination delivers stronger minority-class performance than single-edit variants while remaining lightweight and model agnostic. Third, we benchmark TASER-AE against classical machine learning baselines, prior EHR augmentation methods, and conditional generative baselines such as CVAE and CGAN, and we validate on an independent cohort, demonstrating consistent gains in minority-class detection across adverse events.

We propose TASER-AE (**T**okenized **A**ugmentation for **S**tructured **E**H**R**–**A**dverse **E**vents) a comprehensive pipeline integrating structured data tokenization, targeted NLP-inspired augmentation, and transformer-based classification specifically tailored to AE prediction from EHRs. Rather than processing raw longitudinal time-series, our method operates on patient summaries comprising demographics, prescribed drugs, and recorded complications over a fixed observation window. Our augmentation strategy employs systematic synonym insertion, random token deletion, and controlled shuffling of structured EHR data to produce diverse, high-quality synthetic records for rare-event scenarios. Coupled with transformer-based modeling, TASER-AE is designed to effectively learn complex interaction patterns from both original and augmented data, substantially enhancing model sensitivity to minority-class events while preserving overall accuracy.

## 2 Related Work

### 2.1 Data Augmentation in NLP

Token-level augmentation in the field of NLP has commonly been adapted to enrich text data. Operations include random drop (removing words randomly from a sentence), random swap (swapping two words in a sentence), random synonym insertion (selecting a random word and inserting one of its synonyms), random synonym replacement (replacing a random word with a synonym), and shuffling (permuting the entire word sequence) [34, 44]. These augmentation techniques effectively expand training data, enhancing robustness and performance, especially in low-resource scenarios [6].

### 2.2 Data Augmentation in EHR

Augmenting EHR data borrows ideas from NLP to perturb existing records rather than synthesize them from scratch. Ishikawa et al. [21] replace selected patient-background tokens (patient demographics, drugs, and complications) with semantically similar neighbors learned via a skip-gram model and show gains of up to 40% in F1 on extremely imbalanced AE labels. Choi and Kim [11] propose an order-shuffling strategy that randomly permutes drug, diagnosis, and procedure codes within a visit, improving ROC-AUC by 5.3% for clopidogrel treatment-failure detection, which is a surrogate for AEs related to platelet inhibition. Together with other token-level edits (drop, swap, synonym insertion), these studies demonstrate that lightweight, NLP-inspired transformations can boost minority-class performance without external data.

In parallel, a strand of work has explored fully generative solutions that learn the joint distribution of drug, diagnosis, and demographic codes and draw new synthetic patients from it. Generative-adversarial networks [10, 42] model high-dimensional binary code vectors, and conditional variational auto-encoders [2] extend generation to entire visit sequences. These examples show that EHR augmentation can take many forms, from creating entirely synthetic patient records to permutations or text-based perturbations.

### 2.3 AE Prediction

Predicting treatment-related AEs from clinical data is an active area of research, often employing traditional machine learning or deep learning models. A 2024 systematic review of 59 EHR studies found that decision tree-based algorithms, random forests, and boosting ensembles consistently delivered the best sensitivity–specificity trade-off for diverse drug–event pairs [19]. Few studies report data augmentation within AE investigations. Some studies focus on specific drug-AE problems: for example, Chang et al. [4] built models to predict anthracycline-induced cardiotoxicity in breast cancer patients.

Transformer architectures have been increasingly applied to coded EHR data by treating diagnoses, procedures, and medications as tokens in a sequence. Pre-trained models such as BEHRT [27] and Med-BERT [36] learn context-aware embeddings from large visit corpora and outperform RNN and gradient-boosting baselines on several risk-prediction tasks, while larger foundation models [39] and generative encoder–decoder variants [48] extend these ideas further. These results motivate our use of a transformer encoder for classification; however, rather than relying on large-scale pre-training, we focus on whether targeted data augmentation can deliver comparable minority-class gains with a lightweight, task-specific model trained from scratch.

## 3 Methods

### 3.1 Problem Formulation

We formulate adverse event (AE) prediction as a multi-label classification task enhanced by data augmentation. Let *x* ∈ 𝒱^*T*^ denote a patient representation formed by (i) demographic tokens for age and sex and (ii) the sequence of drug and complication tokens observed within a fixed time window, where 𝒱 is the vocabulary and *T* is the resulting token length. Let *y* ∈ {0, 1}^*C*^ denote the corresponding vector of *C* adverse event labels. Our goal is to learn a mapping *f*_*θ*_ : 𝒱^*T*^→ [0, 1]^*C*^, with trainable parameters *θ*, that generalizes effectively to unseen samples.

To achieve this, we introduce a stochastic augmentation strategy 𝒜 that maps an observed record *x* to semantically equivalent synthetic records 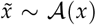. Let 𝒟_pos_ = {(*x, y*) ∈ 𝒟_train_ | ∑_*j*_ *y*_*j*_ *>* 0} denote the subset of training records with at least one adverse event label. We optimize *θ* over the combined distribution of original training records and augmented variants of positive examples.

The desired parameters are:

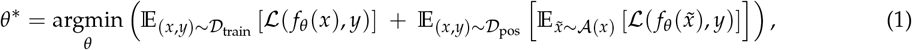

where ℒ is the binary cross-entropy loss. By training on the union of *x* and 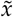, the model preserves the original clinical signal while learning robust feature representations from realistic token-level variations.

### 3.2 Data

We use two EHR cohorts in our study. The first dataset is called Medication Error Avoidance at Regional Scale (MEARS) [12], developed by the Department of Biomedical Informatics at the University of Pittsburgh. This deidentified clinical EHR dataset contains 636,307 patient records from hospitals and outpatient clinics in the University of Pittsburgh Medical Center (UPMC) health system. After receiving an IRB determination that our MEARS analyses do not constitute human subjects research, we sampled 60,610 records for analysis (see Supplementary Material S2 for detailed cohort derivation). Among these, 8,870 instances are labeled with four adverse events (*C* = 4): Falls, Fractures, Gastrointestinal bleed (GI Bleed), and Stroke. We use this dataset for the development and tuning of TASER-AE.

The second dataset is MIMIC-IV [22] from Beth Israel Deaconess Medical Center. For our study, we obtained 144,702 MIMIC-IV patient records, of which 23,951 have at least one of four adverse events. We use this dataset exclusively for independent validation of TASER-AE. No choices are tuned on MIMIC-IV. Further details on the data source, privacy safeguards, and processing are provided in Supplementary Material S3. See Supplementary Figures S1 and S2 for the frequency of each label in MEARS and MIMIC-IV, respectively. The ICD-10 codes used to define these AE labels for both datasets are explained in Supplementary Material S1 and listed in Supplementary Tables.

### 3.3 Patient Representation

As defined above, the patient history is composed of categorical variables. We map these features to integer token IDs in 𝒱. In this study, we do not explicitly model the temporal dimension; specific timestamps and the relative timing of events are disregarded. Instead, we flatten the patient history into a discrete sequence, analogous to a document in natural language processing. Unlike traditional tabular models that require fixed-size feature vectors, this sequence representation *x* = [*w*_1_, …, *w*_*T*_] allows TASER-AE to process variable-length patient backgrounds efficiently via embedding lookups.

### 3.4 Patient Background Thesaurus

To capture semantic relationships between clinical terms, we construct a synonym mapping 𝒮. We first train a Word2Vec [30] model on the corpus of patient sequences in the training set to obtain a dense vector representation **e**_*w*_ ∈ ℝ^*d*^ for every token *w* ∈ 𝒱. We define the semantic similarity between two tokens *w*_*i*_ and *w*_*j*_ using cosine similarity:

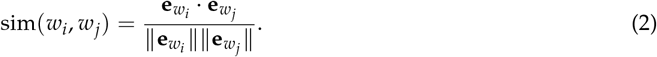

We construct a synonym set 𝒮 (*w*) for each token *w* containing all other terms with high semantic overlap:

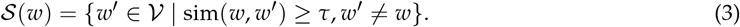

We set the similarity threshold *τ* = 0.8, which is a commonly used cosine-similarity cutoff for identifying close neighbors in embedding spaces [32]. We restrict thesaurus construction to drug and complication terms, excluding demographic tokens. See Figure 1a for a visualization of the similarity dictionary construction.

**Figure 1.**
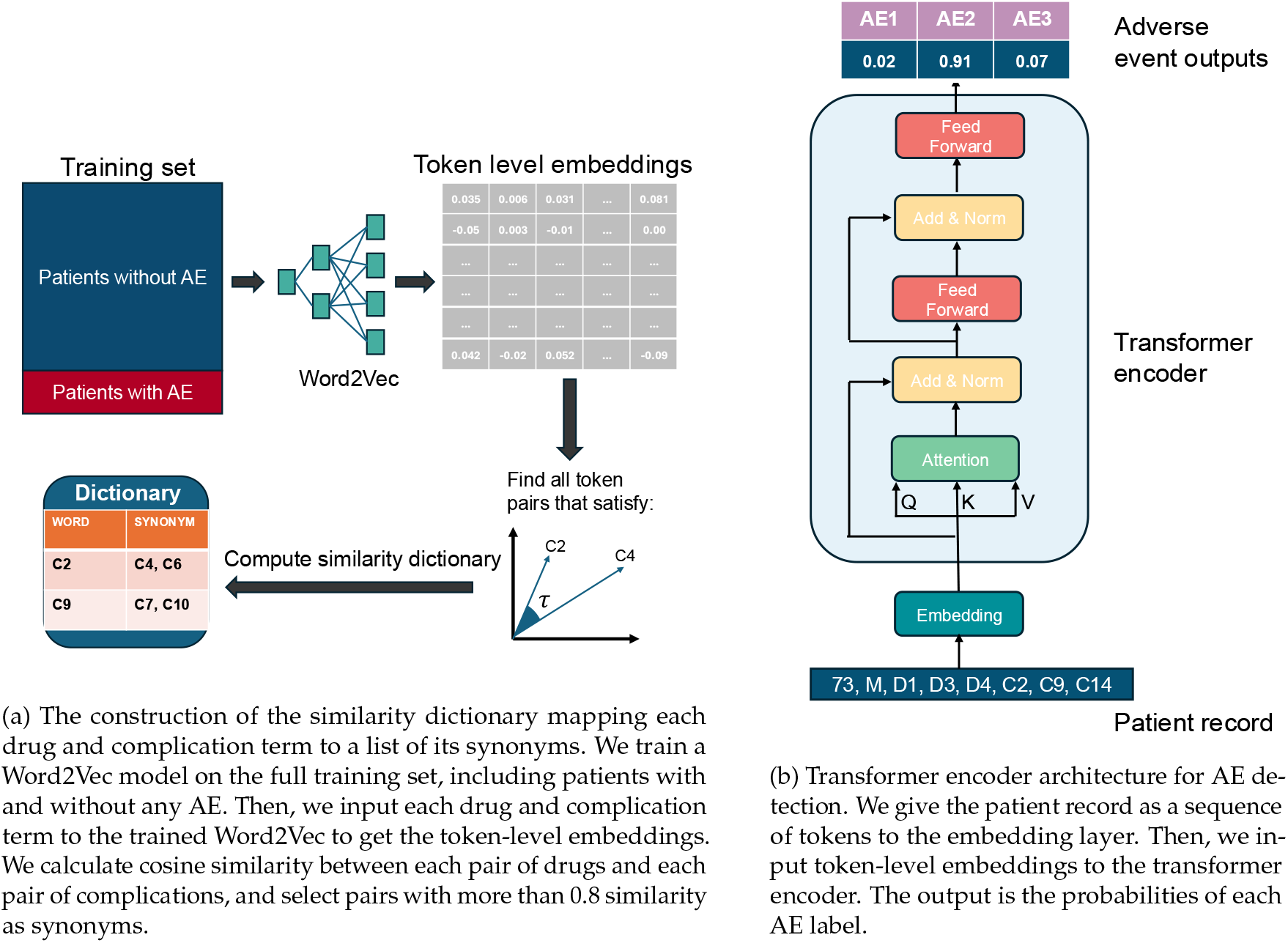
Overview of the proposed system components. (a) illustrates the construction of the similarity dictionary, while (b) shows the Transformer encoder architecture used for AE detection.

### 3.5 TASER-AE’s Adverse Event Detection

We employ a transformer-based encoder for the function *f*_*θ*_. The input sequence *x* is first embedded into a continuous space **X** ∈ ℝ^*T*×*d*^, and we add a learnable positional embedding to each token to capture ordering information. This sequence is then processed by a self-attention mechanism that computes a weighted sum of values based on the compatibility between keys and queries, allowing the model to capture long-range dependencies among clinical features.

The output of the attention block is pooled and passed through a final linear projection layer followed by a sigmoid activation function *σ*(·) to predict the multi-label probabilities:

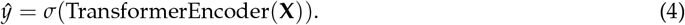

The model is trained to minimize the binary cross-entropy loss:

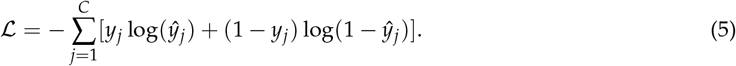

Sequences are padded to a fixed maximum length to facilitate batch processing.

### 3.6 TASER-AE’s Systematic EHR Augmentation

To mitigate class imbalance, we define an augmentation strategy 𝒜 applied specifically to the minority class, patients with recorded adverse events, collectively referred to as positive examples. Let 𝒟_pos_ = {(*x, y*) ∈ 𝒟 | ∑_*j*_ *y*_*j*_ *>* 0} be the set of positive samples. We target a 1× augmentation ratio, meaning we generate |𝒟_pos_| total synthetic samples. To do so, we repeatedly sample a record *x* uniformly at random from 𝒟_pos_ and apply all three transformations in 𝒪 = {*T*_*drop*_, *T*_*shu f*_, *T*_*ins*_} independently, producing three candidate synthetic records per sampled record. Candidates are added to the augmented set until |𝒟_pos_| total synthetic samples have been collected.

These operations are defined as follows for an input sequence *x*:

- **Random Drop (***T*_*drop*_): We simulate missing data by removing a token. Let *i* ∼ 𝒰 { 1, |*x*| }, where 𝒰 denotes the discrete uniform distribution over the integer indices of the sequence. The transformed sequence is:

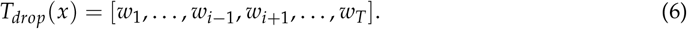

This forces the model to learn robust representations that do not rely on a single specific feature (see Part 1 in Figure 2).
- **Shuffle (***T*_*shu f*_ ): To enforce order-invariance among clinical features (as the order of recorded complications is often arbitrary), we apply a random permutation *π* to the indices of the drug and complication tokens:

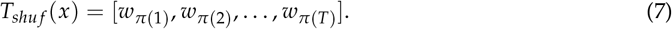

(See Part 2 in Figure 2.)
- **Random Synonym Insertion (***T*_*ins*_): We inject lexical diversity by inserting semantically equivalent terms. We select a token *w*_*i*_ ∈ *x* and identify its synonym set 𝒮 (*w*_*i*_) from the thesaurus. If 𝒮 (*w*_*i*_) ≠ ∅, we sample a synonym *s* ∼ 𝒰 (𝒮 (*w*_*i*_)) and append it to the sequence:

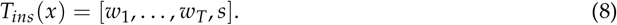

**Figure 2.**
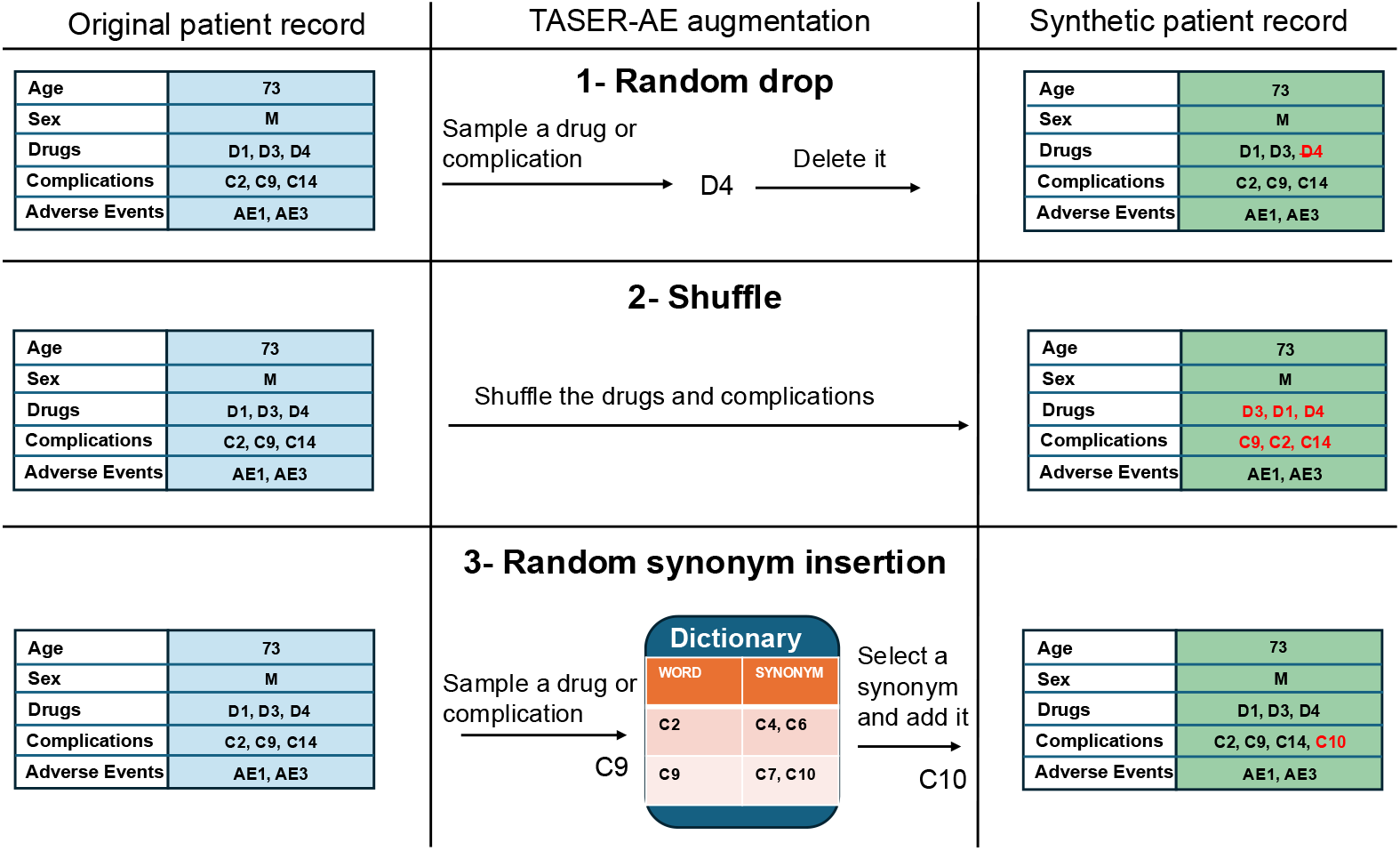
Three operations of TASER-AE augmentation. On left, we show a sample original patient record. We visualize each step for (1) random drop, (2) shuffle, and (3) random synonym insertion operations. On right, the synthetic patient record generated from the operation of that row is shown.

If the selected token has no synonyms, we resample a different token (see Part 3 in Figure 2).

Each synthetic record retains the original label vector *y*. These augmented samples are added to the training set, increasing the effective size and diversity of the positive class.

### 3.7 Baselines

To evaluate our approach, we compare TASER-AE against several baseline augmentation strategies and classification methods.

#### 3.7.1 NLP-style augmentation

In addition to the operations used in TASER-AE (see Section 3.6), we evaluate three other common text augmentation techniques. We define these formally as transformations *T* : 𝒱^*T*^ → 𝒱^*T*^′ applied to a patient sequence *x*.

##### Random swap (*T*_*swap*_)

We select two distinct indices *i, j* from the sequence *x* uniformly at random, such that *w*_*i*_ and *w*_*j*_ belong to the same feature type (e.g., both are drugs or both are complications). The transformation swaps these tokens:

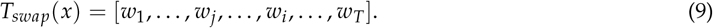

If a feature type contains fewer than two tokens, the operation is skipped or attempted on the alternate feature type. **Random synonym replacement (***T*_*rep*_): We select a token *w*_*i*_ ∈ *x* uniformly at random. We query the thesaurus for 𝒮 (*w*_*i*_). If 𝒮 (*w*_*i*_) ≠ ∅, we sample a synonym *s* ∼ 𝒰 (𝒮 (*w*_*i*_)) and replace the original token:

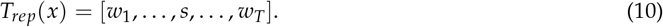

If the sampled synonym *s* already exists in *x*, we retain only one instance to avoid redundancy.

##### Composite insertion (*T*_*comp*_)

This operation inserts multiple tokens simultaneously. We sample one drug token *w*_*d*_ ∈ *x* and one complication token *w*_*c*_ ∈ *x*. For each, we sample a corresponding synonym *s*_*d*_ ∈ 𝒮 (*w*_*d*_) and *s*_*c*_ ∈ 𝒮 (*w*_*c*_). Both synonyms are appended to the sequence:

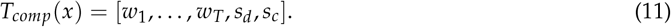

This operation increases the sequence length by up to two tokens, strictly preserving the original tokens while expanding lexical coverage.

#### 3.7.2 Generative AI

We compare against generative models trained to approximate the conditional distribution *p*(*x* | **y**), where **y** ∈ {0, 1}^*C*^ denotes the multi-label AE vector, to synthesize new patient records:

- **Conditional VAE (CVAE):** A variational autoencoder that learns a latent variable model conditioned on the multi-label AE vector *y*.
- **Conditional GAN (CGAN):** A label-conditioned Generative Adversarial Network where the generator *G* (*z, y*) produces synthetic sequences from Gaussian noise *z*.

Detailed architectures, loss functions, and training hyperparameters are provided in Supplementary Material S4.

#### 3.7.3 Classical classifiers

We train three classical machine learning baselines: decision tree, random forest, and XGBoost [7]. Since these models require fixed-size input vectors rather than sequences, we apply a multi-hot encoding function *ϕ* : 𝒱^*T*^ → {0, 1}^|𝒱|^ to the patient records. We tuned all model hyperparameters on the validation split to maximize the macro F1 score. Implementation details and hyperparameter grids appear in Supplementary Material S5.

### 3.8 Study design

We adopt a strict development–validation design to ensure generalizability. Let 𝒟_MEARS_ denote the development cohort and 𝒟_MIMIC_ denote the external validation cohort. All algorithmic design choices, including tokenization strategies, the augmentation set 𝒪, model architecture specifications (e.g., number of layers, embedding dimensions), and optimization hyperparameters, were determined exclusively using 𝒟_MEARS_.

Once the optimal configuration is identified, the methodological protocol is frozen. For external validation, we do not transfer the learned weights; instead, we re-initialize the model parameters *θ* and train *f*_*θ*_ from scratch on 𝒟_MIMIC_ using the exact configuration derived from MEARS. The augmentation logic and model backbone remain identical.

### 3.9 Training

We trained the transformer-based classifier *f*_*θ*_ (Section 3.5) for each experimental scenario. For each tested augmentation strategy, the training set 𝒟_train_ is expanded with synthetic samples. For all experiments, we generate one synthetic sample for every original positive instance (a 1× augmentation ratio). The model parameters *θ* are optimized using Adam [23] (learning rate *α* = 10^−3^, weight decay *λ* = 10^−5^) to minimize the binary cross-entropy loss. These hyperparameters were selected empirically based on preliminary experiments on the MEARS training and validation splits. We employ a fixed stratified split of 𝒟_MEARS_ (64% train, 16% validation, 20% test) to preserve class priors *p*(*y*). We train for a maximum of 100 epochs with early stopping and save the parameter state *θ* corresponding to the epoch with the lowest loss on the validation split.

To evaluate performance on imbalanced data, we calculate Precision, Recall, and F1-score. We report these metrics for each individual adverse event label, as well as their aggregate Macro and Minority averages. For a given metric *M* ∈ {Precision, Recall, F1}, let *M*_*i*_ denote the score for class *i*. The Macro average represents the unweighted mean across all classes:

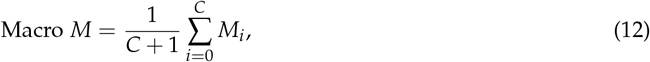

where *C* is the number of adverse event labels and index 0 represents the majority “No Adverse Event” class. Additionally, we compute the *Minority Average*, which restricts the mean to the adverse event classes only:

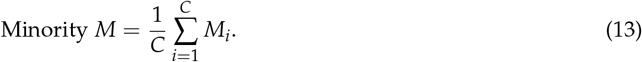

Unlike micro-averaging, these aggregate metrics treat minority classes with equal weight, preventing the majority class from dominating the global evaluation.

## 4 Results

### 4.1 Baselines

#### 4.1.1 Classical Models

We evaluate three classical machine learning classifiers: decision tree, random forest, and XGBoost, trained on the same MEARS features as our neural models. For each classifier, we select the best configuration based on the validation macro F1 and then report its performance on the test set. We describe all implementation details and hyperparameter tuning procedures in Supplementary Material S5. The class-specific performance results of each classifier on MEARS are detailed in Table 2. The results show limited effectiveness for minority class predictions across all classifiers, underscoring the challenge posed by severe class imbalance.

**Table 1.**
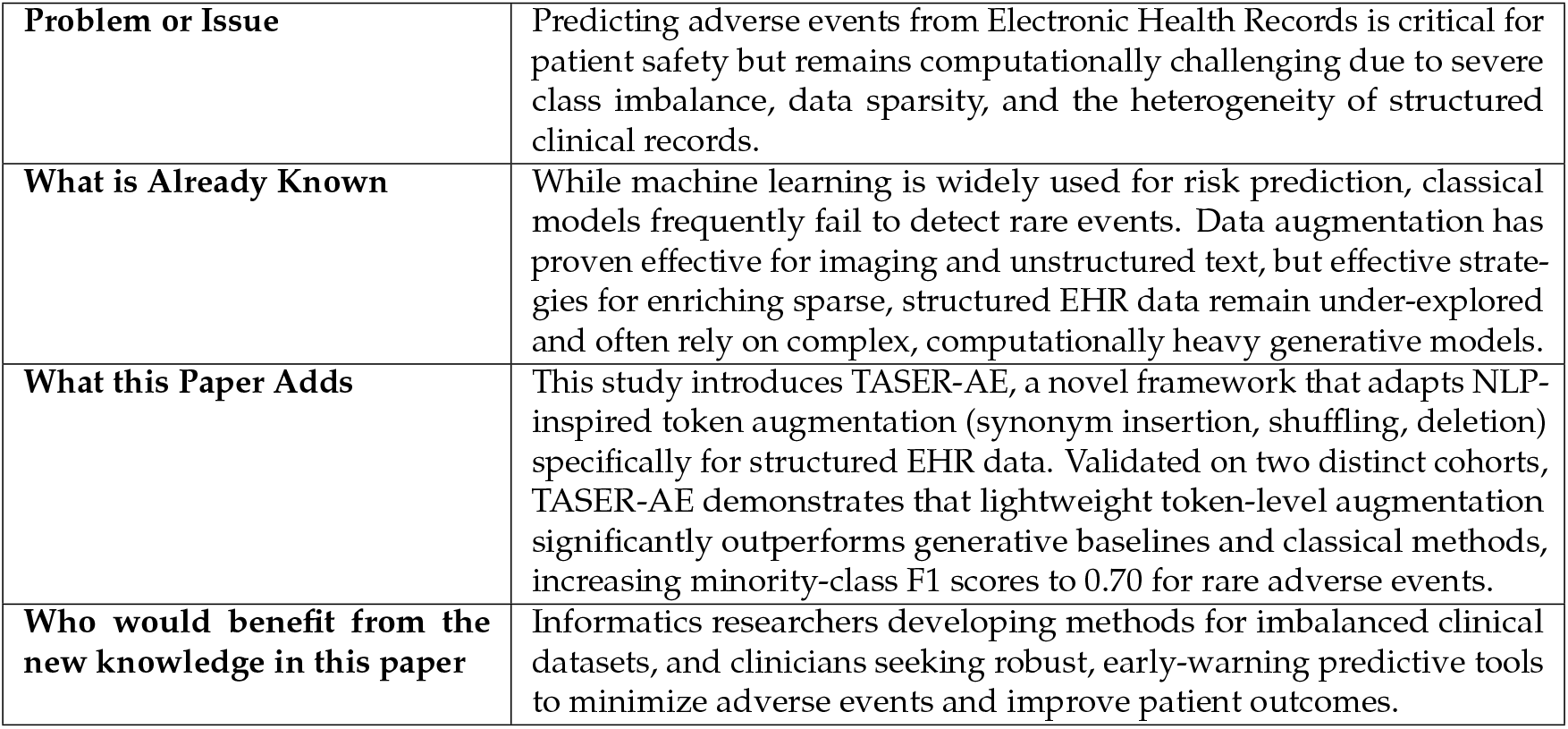
Statement of Significance.

|  |  |
| --- | --- |
| **Problem or Issue** | Predicting adverse events from Electronic Health Records is critical for patient safety but remains computationally challenging due to severe class imbalance, data sparsity, and the heterogeneity of structured clinical records. |
| **What is Already Known** | While machine learning is widely used for risk prediction, classical models frequently fail to detect rare events. Data augmentation has proven effective for imaging and unstructured text, but effective strategies for enriching sparse, structured EHR data remain under-explored and often rely on complex, computationally heavy generative models. |
| **What this Paper Adds** | This study introduces TASER-AE, a novel framework that adapts NLP-inspired token augmentation (synonym insertion, shuffling, deletion) specifically for structured EHR data. Validated on two distinct cohorts, TASER-AE demonstrates that lightweight token-level augmentation significantly outperforms generative baselines and classical methods, increasing minority-class F1 scores to 0.70 for rare adverse events. |
| **Who would benefit from the new knowledge in this paper** | Informatics researchers developing methods for imbalanced clinical datasets, and clinicians seeking robust, early-warning predictive tools to minimize adverse events and improve patient outcomes. |

**Table 2.**
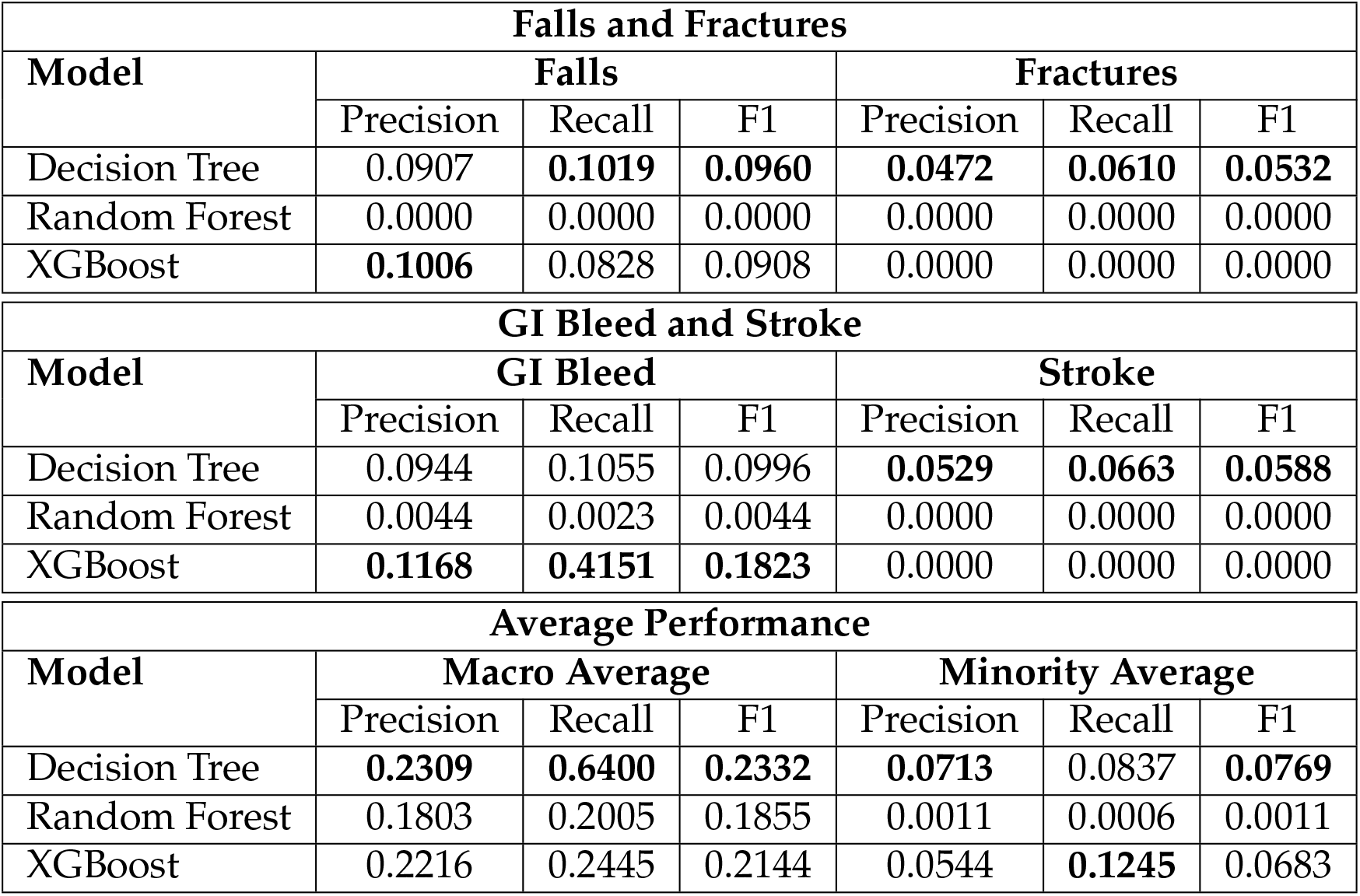
Performance of Classical Models on MEARS Minority Classes.

| Falls and Fractures |  |  |  |  |  |  |
| --- | --- | --- | --- | --- | --- | --- |
| Model | Falls |  |  | Fractures |  |  |
|  | Precision | Recall | F1 | Precision | Recall | F1 |
| Decision Tree | 0.0907 | **0.1019** | **0.0960** | **0.0472** | **0.0610** | **0.0532** |
| Random Forest | 0.0000 | 0.0000 | 0.0000 | 0.0000 | 0.0000 | 0.0000 |
| XGBoost | **0.1006** | 0.0828 | 0.0908 | 0.0000 | 0.0000 | 0.0000 |

| GI Bleed and Stroke |  |  |  |  |  |  |
| --- | --- | --- | --- | --- | --- | --- |
| Model | GI Bleed |  |  | Stroke |  |  |
|  | Precision | Recall | F1 | Precision | Recall | F1 |
| Decision Tree | 0.0944 | 0.1055 | 0.0996 | **0.0529** | **0.0663** | **0.0588** |
| Random Forest | 0.0044 | 0.0023 | 0.0044 | 0.0000 | 0.0000 | 0.0000 |
| XGBoost | **0.1168** | **0.4151** | **0.1823** | 0.0000 | 0.0000 | 0.0000 |

| Average Performance |  |  |  |  |  |  |
| --- | --- | --- | --- | --- | --- | --- |
| Model | Macro Average |  |  | Minority Average |  |  |
|  | Precision | Recall | F1 | Precision | Recall | F1 |
| Decision Tree | **0.2309** | **0.6400** | **0.2332** | **0.0713** | 0.0837 | **0.0769** |
| Random Forest | 0.1803 | 0.2005 | 0.1855 | 0.0011 | 0.0006 | 0.0011 |
| XGBoost | 0.2216 | 0.2445 | 0.2144 | 0.0544 | **0.1245** | 0.0683 |

#### 4.1.2 Transformer on Original Data

We evaluate the transformer classifier on the original (non-augmented) MEARS. The baseline model achieves an F1 of 0.015 (precision 0.171, recall 0.008) for Falls, an F1 of 0.000 for Fractures, an F1 of 0.094 (precision 0.447, recall 0.052) for GI Bleed, and an F1 of 0.041 (precision 0.546, recall 0.021) for Stroke. Consequently, the average minority F1 is extremely low at 0.038, revealing that minority adverse event classes are poorly captured. The baseline model essentially fails to predict the underrepresented outcomes. This behavior is consistent with known results that imbalanced data leads to very low F1 on rare classes unless addressed. This baseline’s poor macro average F1 (0.212) further suggests severe overfitting to the majority class, namely patients without any recorded AE label (the “No adverse event” class).

#### 4.1.3 Transformer on Augmented Data

We apply each augmentation method discussed in Section 3.7.1 individually to the training data and retrain the same transformer classifier. Table 3 reports precision, recall, and F1 for four adverse events on MEARS under several augmentation methods alongside the no-augmentation original data baseline.

**Table 3.** Minority-Class Performance Across Augmentation Methods (MEARS)

| Falls and Fractures |  |  |  |  |  |  |
| --- | --- | --- | --- | --- | --- | --- |
| Method | Falls |  |  | Fractures |  |  |
|  | Precision | Recall | F1 | Precision | Recall | F1 |
| No Augmentation | 0.1707 | 0.0081 | 0.0154 | 0.0000 | 0.0000 | 0.0000 |
| Random Drop | 0.4659 | 0.5127 | 0.4882 | 0.6275 | 0.5818 | 0.6038 |
| Random Swap | 0.3539 | 0.3048 | 0.3275 | 0.5943 | 0.3818 | 0.4649 |
| Random Synonym Repl. | 0.3278 | 0.6848 | 0.4434 | 0.4436 | 0.6909 | 0.5403 |
| Random Synonym Insert. | 0.5570 | 0.5416 | 0.5492 | 0.5781 | 0.6727 | 0.6218 |
| Shuffle | 0.4639 | 0.5855 | 0.5176 | 0.5687 | 0.7273 | 0.6383 |
| Composite Insertion | 0.3893 | 0.2194 | 0.2806 | 0.5179 | 0.5273 | 0.5225 |
| Conditional VAE | 0.3348 | 0.0865 | 0.1375 | 0.5455 | 0.1775 | 0.2679 |
| Conditional GAN | 0.3028 | 0.0966 | 0.1465 | 0.4762 | 0.1183 | 0.1896 |
| Ishikawa et al. | 0.3521 | 0.6351 | 0.4530 | 0.5435 | 0.6061 | 0.5731 |
| **TASER-AE** | **0.6136** | **0.7171** | **0.6613** | **0.6294** | **0.7515** | **0.6851** |

| GI Bleed and Stroke |  |  |  |  |  |  |
| --- | --- | --- | --- | --- | --- | --- |
| Method | GI Bleed |  |  | Stroke |  |  |
|  | Precision | Recall | F1 | Precision | Recall | F1 |
| No Augmentation | 0.4472 | 0.0524 | 0.0939 | 0.5455 | 0.0211 | 0.0407 |
| Random Drop | 0.5464 | 0.4099 | 0.4684 | 0.5975 | 0.3345 | 0.4289 |
| Random Swap | 0.3096 | 0.3603 | 0.3300 | 0.3739 | 0.5739 | 0.4528 |
| Random Synonym Repl. | 0.3465 | 0.6683 | 0.4564 | 0.3891 | 0.7042 | 0.5013 |
| Random Synonym Insert. | 0.5745 | 0.4852 | 0.5261 | 0.6133 | 0.6479 | 0.6301 |
| Shuffle | 0.4100 | 0.5844 | 0.4819 | 0.5491 | 0.6690 | 0.6032 |
| Composite Insertion | 0.3604 | 0.2669 | 0.3067 | 0.3800 | 0.4683 | 0.4196 |
| Conditional VAE | 0.3333 | 0.0575 | 0.0981 | 0.3462 | 0.1111 | 0.1682 |
| Conditional GAN | 0.3280 | 0.0773 | 0.1251 | 0.4018 | 0.1389 | 0.2064 |
| Ishikawa et al. | 0.2988 | 0.6759 | 0.4144 | 0.3949 | **0.7148** | 0.5088 |
| **TASER-AE** | **0.6315** | **0.7073** | **0.6673** | **0.7067** | 0.7042 | **0.7055** |

| Average Performance |  |  |  |  |  |  |
| --- | --- | --- | --- | --- | --- | --- |
| Method | Macro Average |  |  | Minority Average |  |  |
|  | Precision | Recall | F1 | Precision | Recall | F1 |
| No Augmentation | 0.4056 | 0.2091 | 0.2123 | 0.2909 | 0.0204 | 0.0375 |
| Random Drop | 0.6354 | 0.5498 | 0.5828 | 0.5593 | 0.4597 | 0.4973 |
| Random Swap | 0.5094 | 0.4838 | 0.4862 | 0.4079 | 0.4052 | 0.3938 |
| Random Synonym Repl. | 0.4941 | 0.6892 | 0.5502 | 0.3768 | 0.6871 | 0.4854 |
| Random Synonym Insert. | 0.6529 | 0.6543 | 0.6520 | 0.5807 | 0.5869 | 0.5818 |
| Shuffle | 0.5872 | 0.6847 | 0.6280 | 0.4979 | 0.6416 | 0.5603 |
| Composite Insertion | 0.5065 | 0.4766 | 0.4845 | 0.4119 | 0.3705 | 0.3824 |
| Conditional VAE | 0.4728 | 0.2765 | 0.3136 | 0.3900 | 0.1082 | 0.1679 |
| Conditional GAN | 0.3653 | 0.2025 | 0.1866 | 0.3772 | 0.1078 | 0.1669 |
| Ishikawa et al. | 0.5094 | 0.6559 | 0.5444 | 0.3973 | 0.6580 | 0.4873 |
| **TASER-AE** | **0.7104** | **0.7615** | **0.7336** | **0.6453** | **0.7200** | **0.6798** |

Simple token-level edits improve performance. Random drop, random swap, random synonym replacement, random synonym insertion, and shuffle each raise both recall and F1 with varying trade-offs. Methods that increase lexical diversity while preserving clinical content, such as random synonym insertion and shuffle, tend to deliver higher F1 than random swap or composite insertion. For example, random synonym insertion attains a minority average F1 of 0.582, driven by strong individual scores such as a Falls F1 of 0.549 and Fractures F1 of 0.622. Shuffle shows similar gains with a minority average F1 of 0.560. Composite insertion underperforms these methods, sacrificing recall and resulting in a lower minority average F1 of 0.382.

We evaluate CVAE and CGAN models for augmentation. These methods performed very poorly. While they offer a slight improvement over the no-augmentation baseline for some outcomes, they are dramatically outperformed by the simple token-level edits. Their primary failure is extremely low recall across all four adverse events, resulting in minority average F1 scores of only 0.168 (CVAE) and 0.167 (CGAN). This performance, which is only marginally better than the baseline average of 0.038, suggests that these generative models struggled to produce synthetic samples that were sufficiently coherent or diverse to effectively expand the minority class training data.

These results demonstrate a clear pattern: all edit-based augmentation methods drastically improve performance on rare adverse events, at the cost of varying impact on overall accuracy. For example, Falls (baseline F1 0.015) reach F1 0.28–0.55 with augmentation, and Fractures (baseline 0.000) reach 0.52–0.64. Stroke prediction improves from F1 0.041 to over 0.42 in all cases. This is consistent with other studies. The uplift in F1 is similar to previous reports; for instance, Ishikawa et al. [21] found that an NLP-inspired augmentation technique gave 40.0% improvement in the F1-score on rare adverse events, and we see analogous gains here.

### 4.2 TASER-AE

TASER-AE achieves the strongest performance on MEARS across all four adverse events and all three metrics (Table 3). Its minority-class average F1 of 0.680 (precision 0.645, recall 0.720) exceeds the next-best single-edit method, random synonym insertion, by nearly 0.10 in F1 and the prior EHR augmentation baseline of Ishikawa et al. by 0.19. Unlike methods that boost recall at the expense of precision—Ishikawa et al. achieve high recall (e.g., 0.715 for Stroke) but substantially lower precision (0.395)—TASER-AE raises both metrics simultaneously. This pattern holds across all four outcomes: for every adverse event, TASER-AE’s F1 exceeds that of every competing method by at least 0.06. The simultaneous gains in precision and recall indicate that the combined augmentation strategy produces diverse yet clinically coherent synthetic records rather than simply inflating the positive class with noisy samples.

### 4.3 Validation on an Independent Dataset

We evaluate the frozen TASER-AE pipeline on MIMIC-IV as an independent validation cohort. No model selection, hyperparameter tuning, augmentation choice, or thresholding is based on MIMIC-IV; we initialize a new model and train it from scratch using the protocol established on MEARS.

Classical models on MIMIC-IV achieve near-zero minority-class F1 (Supplementary Table S1), with the best performer, Decision Tree, reaching only 0.098. The no-augmentation transformer performs somewhat better but still yields a minority average F1 of only 0.219, driven by high precision but very low recall (Table 4).

**Table 4.** Minority-Class Performance Across Augmentation Methods (MIMIC-IV)

| Falls and Fractures |  |  |  |  |  |  |
| --- | --- | --- | --- | --- | --- | --- |
| Method | Falls |  |  | Fractures |  |  |
|  | Precision | Recall | F1 | Precision | Recall | F1 |
| No Augmentation | 0.3333 | 0.0948 | 0.1476 | 0.2925 | 0.0475 | 0.0818 |
| Random Drop | 0.4671 | 0.2245 | 0.3033 | 0.6377 | 0.1350 | 0.2228 |
| Random Swap | 0.4820 | 0.2228 | 0.3048 | **0.7304** | 0.1288 | 0.2190 |
| Random Synonym Repl. | 0.4925 | 0.0915 | 0.1543 | 0.4737 | 0.0552 | 0.0989 |
| Random Synonym Insert. | 0.4677 | 0.2406 | 0.3177 | 0.6908 | 0.1610 | 0.2612 |
| Shuffle | 0.4462 | 0.1907 | 0.2672 | 0.6259 | 0.1411 | 0.2303 |
| Composite Insertion | 0.4614 | **0.2816** | **0.3497** | 0.6440 | 0.1887 | 0.2918 |
| Conditional VAE | **0.5021** | 0.1303 | 0.2069 | 0.5714 | 0.1350 | 0.2184 |
| Conditional GAN | 0.4325 | 0.2007 | 0.2741 | 0.4818 | 0.0813 | 0.1391 |
| Ishikawa et al. | 0.4607 | 0.2567 | 0.3297 | 0.5237 | 0.2546 | 0.3426 |
| **TASER-AE** | 0.4564 | 0.2700 | 0.3393 | 0.5054 | **0.2853** | **0.3647** |

| GI Bleed and Stroke |  |  |  |  |  |  |
| --- | --- | --- | --- | --- | --- | --- |
| Method | GI Bleed |  |  | Stroke |  |  |
|  | Precision | Recall | F1 | Precision | Recall | F1 |
| No Augmentation | **0.6400** | 0.3656 | 0.4653 | 0.4291 | 0.1161 | 0.1827 |
| Random Drop | 0.5871 | 0.4598 | 0.5157 | 0.6308 | **0.5367** | 0.5800 |
| Random Swap | 0.5872 | 0.4411 | 0.5038 | 0.6302 | 0.5208 | 0.5703 |
| Random Synonym Repl. | 0.5746 | 0.4500 | 0.5047 | 0.6434 | 0.5016 | 0.5637 |
| Random Synonym Insert. | 0.6097 | 0.3994 | 0.4827 | 0.6998 | 0.4345 | 0.5361 |
| Shuffle | 0.5484 | **0.4951** | **0.5204** | 0.6107 | 0.5229 | 0.5634 |
| Composite Insertion | 0.5537 | 0.4652 | 0.5056 | **0.7335** | 0.4366 | 0.5474 |
| Conditional VAE | 0.6030 | 0.3808 | 0.4668 | 0.7302 | 0.5016 | 0.5947 |
| Conditional GAN | 0.5439 | 0.3705 | 0.4407 | 0.6739 | 0.4622 | 0.5483 |
| Ishikawa et al. | 0.5983 | 0.4092 | 0.4860 | 0.6719 | 0.5037 | 0.5758 |
| **TASER-AE** | 0.6040 | 0.4132 | 0.4907 | 0.6943 | 0.5346 | **0.6041** |

| Averages |  |  |  |  |  |  |
| --- | --- | --- | --- | --- | --- | --- |
| Method | Macro Average |  |  | Minority Average |  |  |
|  | Precision | Recall | F1 | Precision | Recall | F1 |
| No Augmentation | 0.5174 | 0.3151 | 0.3597 | 0.4237 | 0.1560 | 0.2194 |
| Random Drop | 0.6471 | 0.4524 | 0.5063 | 0.5807 | 0.3390 | 0.4054 |
| Random Swap | **0.6681** | 0.4449 | 0.5017 | **0.6075** | 0.3284 | 0.3995 |
| Random Synonym Repl. | 0.6199 | 0.3979 | 0.4449 | 0.5461 | 0.2746 | 0.3304 |
| Random Synonym Insert. | 0.6765 | 0.4251 | 0.4999 | 0.6170 | 0.3089 | 0.3994 |
| Shuffle | 0.6304 | 0.4459 | 0.4962 | 0.5578 | 0.3374 | 0.3953 |
| Composite Insertion | 0.6617 | 0.4509 | 0.5186 | 0.5981 | 0.3430 | 0.4236 |
| Conditional VAE | 0.6598 | 0.4196 | 0.4814 | 0.6017 | 0.2869 | 0.3717 |
| Conditional GAN | 0.6044 | 0.4097 | 0.4628 | 0.5330 | 0.2787 | 0.3506 |
| Ishikawa et al. | 0.6350 | 0.4601 | 0.5264 | 0.5636 | 0.3561 | 0.4335 |
| **TASER-AE** | 0.6369 | **0.4751** | **0.5393** | 0.5651 | **0.3757** | **0.4497** |

Among augmentation methods, simple token-level edits consistently outperform the generative baselines (CVAE and CGAN), which are characterized by low recall and minority average F1 scores below 0.38. While individual edit methods and Ishikawa et al. each lead on specific outcomes, for example, Composite Insertion achieves the highest Falls F1 (0.350) and Shuffle the highest GI Bleed F1 (0.520), none is uniformly dominant. TASER-AE proves to be the most robust across the AEs, achieving the highest F1 for Fractures (0.365) and Stroke (0.604) and the strongest aggregate scores: a macro average F1 of 0.539 and a minority average F1 of 0.450. These gains reflect substantially higher recall at competitive precision, indicating better coverage of true cases rather than overfitting to easy positives.

The frozen augmentation and modeling choices derived on MEARS transfer to a distinct institution and coding distribution, yielding consistent minority-class improvements without any MIMIC-specific tuning. This external validation supports TASER-AE’s robustness under dataset shift and its practical utility for AE detection in new clinical settings.

### 4.4 Visualization of Augmented Data

We visualize real and augmented samples using t-SNE embeddings for both the MEARS (Figure 3 top row) and MIMIC-IV (Figure 3 bottom row) datasets to qualitatively assess the distribution and coverage provided by our augmentation approach. For both datasets, we randomly select 50% of the samples to improve visualization clarity, resulting in 4,180 points for the MEARS real and augmented subplots (Figure 3(a, b)) and 8,360 points for the combined plot (Figure 3(c)) and 3,389 points for the MIMIC real and augmented subplots (Figure 3(d, e)) and 6,778 points in the combined plot (Figure 3(f)).

**Figure 3.**
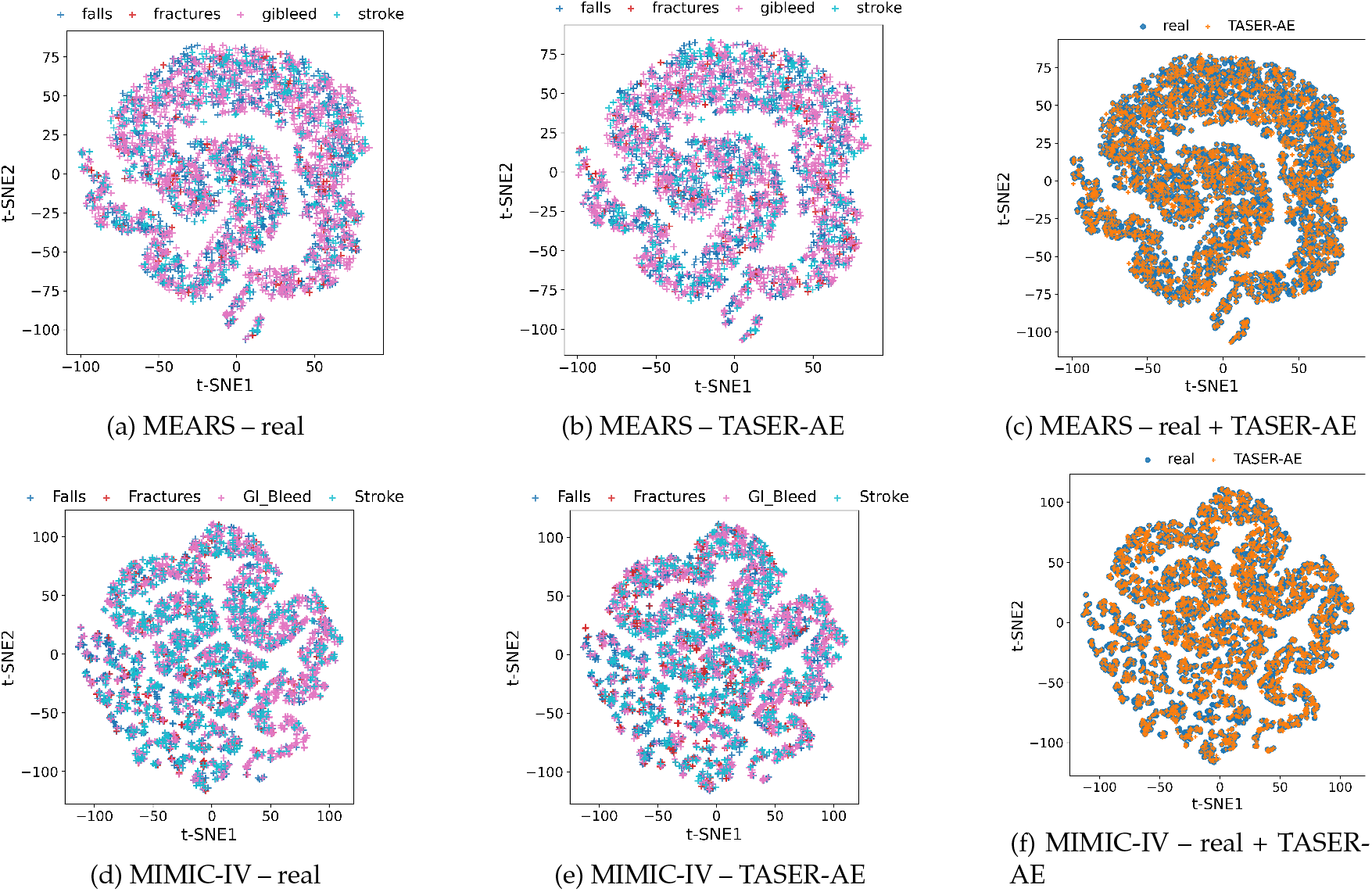
t-SNE projections of real and augmented EHR samples for MEARS (top row) and MIMIC-IV (bottom row). Augmented points (TASER-AE) tend to fill gaps in the manifold rather than form outliers, increasing overall coverage.

Figure 3(a) displays real samples categorized by adverse events, with distinct colors representing each class. Figure 3(b) shows augmented samples generated by our proposed TASER-AE method, similarly color-coded by adverse events. Figure 3(c) combines both real and augmented samples, illustrating the relationship between the original and synthetic data. For MIMIC-IV, Figure 3(d) represents real adverse event samples, Figure 3(e) illustrates TASER-AE augmented samples, and Figure 3(f) combines both.

In these visualizations, augmented samples consistently fill spatial gaps between real samples and never appear isolated or distant from authentic data points. This indicates that our augmentation approach effectively enriches data representation without introducing unrealistic or out-of-distribution samples. The augmented data enhances coverage, suggesting increased robustness and generalization capabilities for predictive models trained on these enriched datasets.

### 4.5 Effect of Synthetic Sample Count

The model attains its highest macro-averaged F1 score when the number of synthetic samples is equal to that of the original dataset (1×), thereby forming a training set with approximately equal number of positive and negative examples (Figure 4a and 4b). This configuration provides an adequate data size while preserving the realism of the overall data distribution. When only a small proportion of synthetic data is incorporated, model performance is constrained by the limited representation of minority-labeled samples. In contrast, when the proportion of synthetic data exceeds 1×, the macro-averaged F1 score declines, likely due to a distributional shift introduced by the excessive inclusion of synthetic samples. These results indicate that a balanced integration of real and synthetic data yields the most effective model performance.

**Figure 4.**
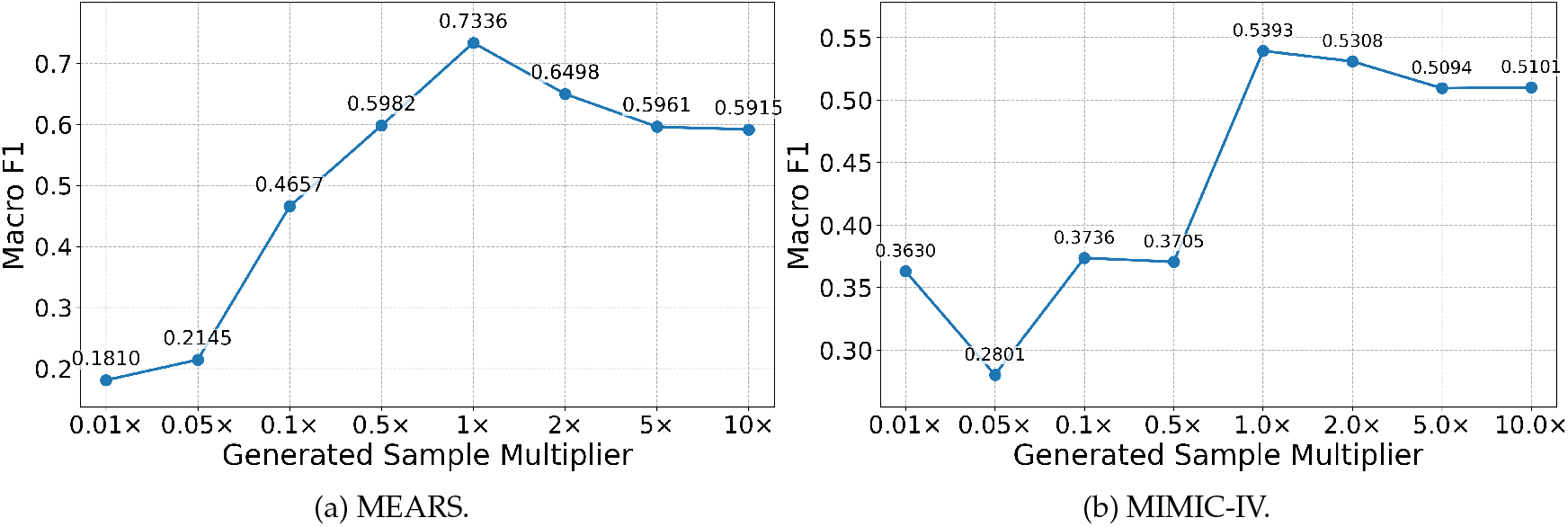
Macro-averaged F1 score versus number of generated samples in both datasets. Macro F1 is computed over the adverse event labels only and excludes the majority “No adverse event” class.

## 5 Discussion

In this study, we demonstrate that simple, targeted data-augmentation strategies can markedly improve the prediction of drug-related AEs from EHR data, specifically prescribed drugs and assigned conditions and very basic demographics. Our proposed method, combining random synonym insertion, random drop, and shuffling, achieves minority-class F1 scores up to 0.70 (an absolute gain of more than 0.64), substantially outperforming both the unaugmented transformer baseline and prior EHR-specific augmentation studies. These gains translate directly into more reliable identification of at-risk patients.

Clinically, our approach has three key implications. First, by improving the detection of patients at risk for a drug-related AE, the model can provide real-time feedback to clinicians at the point of drug ordering or administration, potentially flagging high-risk combinations before harm occurs. Second, alerts based on continuous EHR monitoring could trigger timely interventions (e.g., dose adjustment, additional labs, or closer nursing observation), thereby mitigating the severity of emerging AEs. Third, at scale, such preemptive risk stratification has the potential to reduce overall AE incidence, improving patient safety and lowering healthcare costs associated with prolonged hospitalizations or complications.

Despite these promising findings, several limitations warrant consideration. We developed and tuned the augmentation pipeline on one dataset (MEARS) and validated it on another (MIMIC-IV). In practice, dataset shift remains a challenge; differences in coding systems, local conventions, and institution-level workflows mean that directly transferring trained models or augmentation dictionaries may not yield comparable performance without local retraining. Furthermore, our study focuses on structured text features (drugs and complications) and does not yet incorporate other EHR modalities such as vitals, laboratory trajectories, or free-text clinical notes, which may provide complementary early signals of emerging adverse events.

Future work will focus on addressing these heterogeneity and modality challenges. To overcome domain shift, techniques such as adversarial feature alignment or meta-learning could be employed to learn augmentation strategies that generalize across institutions. We also aim to extend TASER-AE to leverage the temporal structure of longitudinal EHR data, as time-stamped sequences of events better reflect how clinicians perceive evolving risk. Additionally, given that data imbalance can disproportionately affect underrepresented patient groups, it is critical to evaluate whether augmentation improves or worsens fairness, ensuring that predictive gains are equitable across demographic subgroups. Finally, moving beyond stochastic perturbations to causal-inference–driven augmentation could generate synthetic examples that reflect realistic “what if” scenarios, further enhancing the utility of the model for clinical decision support.

## 6 Conclusion

Predicting adverse events from EHR is a critical component of patient safety, yet it remains computationally challenging due to the extreme rarity of these events and the heterogeneity of structured clinical data. In this study, we presented TASER-AE, a novel framework that adapts NLP-inspired token augmentation techniques to enrich EHR datasets.

Our results demonstrate that semantically aware augmentation strategies substantially outperform both traditional machine learning classifiers and computationally intensive generative baselines. By populating the minority class with clinically coherent synthetic records, TASER-AE achieved substantial improvements in minority-class sensitivity and F1 scores. Furthermore, the successful validation of our approach on the independent MIMIC-IV cohort underscores the method’s robustness and potential for generalization.

Ultimately, this work establishes that complex generative methods are not always necessary for effective EHR augmentation; rather, targeted, domain-informed token manipulation can yield superior predictive performance. By enabling more accurate identification of high-risk patients, TASER-AE offers a practical pathway toward robust clinical decision support, allowing for timely interventions that minimize harm and improve patient outcomes.

## Data Availability

The MEARS dataset was used under a data use agreement with the University of Pittsburgh Medical Center and the Department of Aging, Commonwealth of Pennsylvania, and is not publicly available due to privacy and contractual restrictions. The MIMIC-IV dataset is available to credentialed researchers through PhysioNet (https://physionet.org/content/mimiciv/) upon completion of required training and application.

https://physionet.org/content/mimiciv/3.1/

## Acknowledgments

This work was supported in part by Medication Error Avoidance at Region Scale (MEARS), Jewish Health-care Foundation, Pittsburgh, PA; University of Pittsburgh School of Public Health; US National Science Foundation [III-2232121] and the US National Institutes of Health [R01HG012470]. Data extraction provided by Health Record Research Request (R3) service, Department of Biomedical Informatics, School of Medicine, University of Pittsburgh, sponsored in part by the Clinical and Translational Sciences Institute and Institute for Precision Medicine, University of Pittsburgh, and University of Pittsburgh Medical Center; Neptune Data Warehouse (NCATS, NIH, UL1 TR001857). Data use agreement through the Department of Aging, Commonwealth of Pennsylvania. The sponsors had no role in the preparation or review of this research.

## References

[1] Ashfaq, A. Sant’Anna, M. Lingman, and S. Nowaczyk. Readmission prediction using deep learning on electronic health records. Journal of Biomedical Informatics, 97:103256, 2019.

[2] S. Biswal, S. Ghosh, J. Duke, B. Malin, W. Stewart, C. Xiao, and J. Sun. EVA: Generating longitudinal electronic health records using conditional variational autoencoders. In Machine Learning for Healthcare Conference, pages 260–282. PMLR, 2021.

[3] Centers for Medicare & Medicaid Services. Electronic Health Records. https://www.cms.gov/priorities/key-initiatives/e-health/records, 2025. Accessed 14 July 2025.

[4] W.-T. Chang, C.-F. Liu, Y.-H. Feng, C.-T. Liao, J.-J. Wang, Z.-C. Chen, H.-C. Lee, and J.-Y. Shih. An artificial intelligence approach for predicting cardiotoxicity in breast cancer patients receiving anthracycline. Archives of Toxicology, 96(10):2731–2737, 2022.

[5] H. Chen, L. Dan, Y. Lu, M. Chen, and J. Zhang. An improved data augmentation approach and its application in medical named entity recognition. BMC Medical Informatics and Decision Making, 24(1):221, 2024.

[6] J. Chen, D. Tam, C. Raffel, M. Bansal, and D. Yang. An empirical survey of data augmentation for limited data learning in NLP. Transactions of the Association for Computational Linguistics, 11:191–211, 2023.

[7] T. Chen and C. Guestrin. XGBoost: A Scalable Tree Boosting System. arXiv, Mar. 2016. doi: 10.1145/2939672.2939785.

[8] X. Chen and Y. Du. Enhancing medical text classification with GAN-based data augmentation and multi-task learning in BERT. Scientific Reports, 15(1):13854, 2025.

[9] P. Chlap, H. Min, N. Vandenberg, J. Dowling, L. Holloway, and A. Haworth. A review of medical image data augmentation techniques for deep learning applications. Journal of Medical Imaging and Radiation Oncology, 65(5):545–563, 2021.

[10] E. Choi, S. Biswal, B. Malin, J. Duke, W. F. Stewart, and J. Sun. Generating multi-label discrete patient records using generative adversarial networks. In Machine Learning for Healthcare Conference, pages 286–305. PMLR, 2017.

[11] S. Choi and S. Kim. Data augmentation method for modeling health records with applications to clopidogrel treatment failure detection. arXiv preprint 2402.18046, 2024.

[12] Department of Biomedical Informatics, University of Pittsburgh. Projects. https://www.dbmi.pitt.edu/projects/, 2025. Accessed 14 July 2025.

[13] V. S. Dsouza, L. Leyens, J. R. Kurian, A. Brand, and H. Brand. Artificial intelligence in pharmacovigilance: A systematic review on predicting adverse drug reactions in hospitalized patients. Research in Social and Administrative Pharmacy, 2025.

[14] M. Ducharme and L. Boothby. Analysis of adverse drug reactions for preventability. International Journal of Clinical Practice, 61(1):157–161, 2007.

[15] Feng, D. Le, and A. B. McCoy. Using electronic health records to identify adverse drug events in ambulatory care: a systematic review. Applied Clinical Informatics, 10(01):123–128, 2019.

[16] F. Garcea, A. Serra, F. Lamberti, and L. Morra. Data augmentation for medical imaging: A systematic literature review. Computers in Biology and Medicine, 152:106391, 2023.

[17] S. Gholampour. Impact of nature of medical data on machine and deep learning for imbalanced datasets: clinical validity of SMOTE is questionable. Machine Learning and Knowledge Extraction, 6(2):827–841, 2024.

[18] M. Hohl, S. S. Small, D. Peddie, K. Badke, C. Bailey, and E. Balka. Why clinicians don’t report adverse drug events: qualitative study. JMIR Public Health and Surveillance, 4(1):e9282, 2018.

[19] Q. Hu, Y. Chen, D. Zou, Z. He, and T. Xu. Predicting adverse drug event using machine learning based on electronic health records: a systematic review and meta-analysis. Frontiers in Pharmacology, 15:1497397, 2024.

[20] Q. Hu, J. Li, X. Li, D. Zou, T. Xu, and Z. He. Machine learning to predict adverse drug events based on electronic health records: a systematic review and meta-analysis. Journal of International Medical Research, 52(12):03000605241302304, 2024.

[21] T. Ishikawa, T. Yakoh, and H. Urushihara. An NLP-inspired data augmentation method for adverse event prediction using an imbalanced healthcare dataset. IEEE Access, 10:81166–81176, 2022.

[22] E. W. Johnson, L. Bulgarelli, L. Shen, A. Gayles, A. Shammout, S. Horng, T. J. Pollard, S. Hao, Moody, B. Gow, L.-W. H. Lehman, L. A. Celi, and R. G. Mark. MIMIC-IV, a freely accessible electronic health record dataset. Scientific Data, 10(1):1, Jan. 2023.

[23] P. Kingma and J. Ba. Adam: A method for stochastic optimization. arXiv, Dec. 2014. doi: 10.48550/arXiv.1412.6980.

[24] O. Kwon, W. Na, H. Kang, T. J. Jun, J. Kweon, G.-M. Park, Y. Cho, C. Hur, J. Chae, D.-Y. Kang, et al. Electronic medical record–based machine learning approach to predict the risk of 30-day adverse cardiac events after invasive coronary treatment: Machine learning model development and validation. JMIR Medical Informatics, 10(5):e26801, 2022.

[25] L. Landow. Monitoring adverse drug events: the Food and Drug Administration MedWatch reporting system. Regional Anesthesia and Pain Medicine, 23(6):190, 1998.

[26] J. Lazarou, B. H. Pomeranz, and P. N. Corey. Incidence of adverse drug reactions in hospitalized patients: a meta-analysis of prospective studies. JAMA, 279(15):1200–1205, 1998.

[27] Y. Li, S. Rao, J. R. A. Solares, A. Hassaine, R. Ramakrishnan, D. Canoy, Y. Zhu, K. Rahimi, and G. Salimi-Khorshidi. BEHRT: Transformer for electronic health records. Scientific Reports, 10(1):7155, 2020.

[28] M. E. Matheny, I. Ricket, C. A. Goodrich, R. U. Shah, M. E. Stabler, A. M. Perkins, C. Dorn, J. Denton, E. Bray, R. Gouripeddi, et al. Development of electronic health record–based prediction models for 30-day readmission risk among patients hospitalized for acute myocardial infarction. JAMA Network Open, 4(1):e2035782–e2035782, 2021.

[29] P. J. McDonnell and M. R. Jacobs. Hospital admissions resulting from preventable adverse drug reactions. Annals of Pharmacotherapy, 36(9):1331–1336, 2002.

[30] T. Mikolov, K. Chen, G. Corrado, and J. Dean. Efficient estimation of word representations in vector space. arXiv, Jan. 2013. doi: 10.48550/arXiv.1301.3781.

[31] R. Miotto, F. Wang, S. Wang, X. Jiang, and J. T. Dudley. Deep learning for healthcare: review, opportunities and challenges. Briefings in Bioinformatics, 19(6):1236–1246, 2018.

[32] J. Morris, E. Lifland, J. Y. Yoo, J. Grigsby, D. Jin, and Y. Qi. TextAttack: A framework for adversarial attacks, data augmentation, and adversarial training in NLP. In Proceedings of the 2020 Conference on Empirical Methods in Natural Language Processing (EMNLP): System Demonstrations, pages 119–126. Association for Computational Linguistics, 2020.

[33] National Institutes of Health. Adverse Events. https://www.ncbi.nlm.nih.gov/books/NBK558963/, 2025. Accessed 14 July 2025.

[34] M. Rahman, W. Yin, and G. Wang. Data augmentation for text classification with ease. In Proceedings of the 6th International Conference on Natural Language and Speech Processing (ICNLSP 2023), pages 324–332, 2023.

[35] N. Rank, B. Pfahringer, J. Kempfert, C. Stamm, T. Kühne, F. Schoenrath, V. Falk, C. Eickhoff, and Meyer. Deep-learning-based real-time prediction of acute kidney injury outperforms human predictive performance. npj Digital Medicine, 3(1):139, 2020.

[36] L. Rasmy, Y. Xiang, Z. Xie, C. Tao, and D. Zhi. Med-BERT: Pretrained contextualized embeddings on large-scale structured electronic health records for disease prediction. npj Digital Medicine, 4(1):86, 2021.

[37] Shorten and T. M. Khoshgoftaar. A survey on image data augmentation for deep learning. Journal of Big Data, 6(1):1–48, 2019.

[38] J. Soyer, D. Necsoiu, I. Desjardins, D. Lebel, and J.-F. Bussiéres. Identification of discrepancies between adverse drug reactions coded by medical records technicians and those reported by the pharmacovigilance team in pediatrics: An intervention to improve identification, reporting, and coding. Archives de Pédiatrie, 26(7):400–406, 2019.

[39] Steinberg, K. Jung, J. A. Fries, C. K. Corbin, S. R. Pfohl, and N. H. Shah. Language models are an effective representation learning technique for electronic health record data. Journal of Biomedical Informatics, 113:103637, 2021.

[40] N. Tomašev, X. Glorot, J. W. Rae, M. Zielinski, H. Askham, A. Saraiva, A. Mottram, C. Meyer, S. Ravuri, Protsyuk, et al. A clinically applicable approach to continuous prediction of future acute kidney injury. Nature, 572(7767):116–119, 2019.

[41] N. Tomašev, N. Harris, S. Baur, A. Mottram, X. Glorot, J. W. Rae, M. Zielinski, H. Askham, A. Saraiva, V. Magliulo, et al. Use of deep learning to develop continuous-risk models for adverse event prediction from electronic health records. Nature Protocols, 16(6):2765–2787, 2021.

[42] Torfi and E. A. Fox. CorGAN: Correlation-capturing convolutional generative adversarial networks for generating synthetic healthcare records. In FLAIRS Conference, pages 335–340, 2020.

[43] U.S. Department of Health and Human Services Office of Inspector General. Adverse Events. https://oig.hhs.gov/reports/featured/adverse-events/, 2025. Accessed 14 July 2025.

[44] Wei and K. Zou. EDA: Easy data augmentation techniques for boosting performance on text classification tasks. arXiv preprint 1901.11196, 2019.

[45] S. Wittlinger, I. C. Wiest, M. J. Ladani, J. N. Kather, M. P. Ebert, F. Siegel, and S. Belle. How machine learning on real world clinical data improves adverse event recording for endoscopy. npj Digital Medicine, 8(1):424, 2025.

[46] World Health Organization. Patient safety, Sept. 2023. URL https://www.who.int/news-room/fact-sheets/detail/patient-safety.

[47] Xiao, E. Choi, and J. Sun. Opportunities and challenges in developing deep learning models using electronic health records data: a systematic review. Journal of the American Medical Informatics Association, 25(10):1419–1428, 2018.

[48] Z. Yang, A. Mitra, W. Liu, D. Berlowitz, and H. Yu. TransformEHR: Transformer-based encoder-decoder generative model to enhance prediction of disease outcomes using electronic health records. Nature Communications, 14(1):7857, 2023.

[49] Y. Zhao, Z. S.-Y. Wong, and K. L. Tsui. A framework of rebalancing imbalanced healthcare data for rare events’ classification: a case of look-alike sound-alike mix-up incident detection. Journal of Healthcare Engineering, 2018(1):6275435, 2018.

[50] Zhu, S. Pu, J. He, D. Su, W. Cai, X. Xu, and H. Liu. Processing imbalanced medical data at the data level with assisted-reproduction data as an example. BioData Mining, 17(1):29, 2024.

