## Supplementary Material for "Augmenting Electronic Health Records for Adverse Event Detection"

Gün Kaynar[1], Zhaoyi You[1], Richard D. Boyce[2], Takahiro Yakoh[3], Carl Kingsford[1]
[1]Ray and Stephanie Lane Computational Biology Department, School of Computer Science, Carnegie Mellon University
[2]Department of Biomedical Informatics, University of Pittsburgh
[3]Department of System Design Engineering, Keio University, Yokohama, Japan

February 10, 2026

#### S1. AE code lists

Adverse Event (AE) labels for the MEARS cohort are derived using the ICD-10-CM root codes listed in Supplementary Tables S2–S5 (Falls, Fractures, GI Bleed, and Stroke). Since MEARS data is normalized to the OMOP Common Data Model (CDM), conditions are primarily stored as SNOMED concept IDs. We map the target ICD-10 definitions to their corresponding SNOMED equivalents using the standard concept relationships provided by the OMOP vocabulary service.

For the MIMIC-IV cohort, we apply the identical adverse event definitions and utilize the same ICD-10-CM root codes listed in Supplementary Tables S2–S5 (Falls, Fractures, GI Bleed, and Stroke). To ensure complete coverage within the MIMIC-IV schema, we normalize the recorded diagnosis codes (removing periods and capitalizing) and apply a prefix-matching strategy to capture all specific descendants of the target root codes (e.g., matching `K25` captures `K25.x`). Detailed expansion logic is provided in Section S3.

#### S2. Data processing and label derivation for MEARS

##### Cohort Construction

The MEARS dataset aggregates de-identified inpatient and outpatient Electronic Health Records (EHR) from the University of Pittsburgh Medical Center (UPMC) covering the period from January 1, 2015, to December 31, 2018. We identified a core cohort of participants in Pennsylvania's Pharmaceutical Assistance Contract for the Elderly (PACE/PACENET) program [6] who had UPMC encounters during this window. PACE constitutes a curated pharmaceutical assistance program aimed at optimizing prescribing practices for older adults. To reduce selection bias, this core group was complemented by a random sample of non-PACE UPMC patients from the same timeframe, matched 5:1 on age, biological sex, and financial coverage status.

##### Standardization

Data was extracted into a relational database conforming to the OMOP Common Data Model [5] version 5.3. All clinical concepts (conditions, drug exposures, procedures, and measurements) were mapped to standard vocabularies (SNOMED-CT for conditions, RxNorm for medications) in accordance with OMOP specifications.

##### Phenotype Definitions

We used SQL queries and the Atlas platform [4] to define AE cohorts.

- **Stroke:** We used ICD-10 codes for Ischemic and Hemorrhagic Stroke, aligning with the Cipher EHR phenotype library [7]. Validation studies on Medicare claims indicate this phenotype achieves a sensitivity of 94.5% (95% CI: 88.5–98.0%) and specificity of 98.4% (95% CI: 98.0–98.8%) [2].

- **GI Bleed:** We selected ICD-10 codes for gastrointestinal or peptic ulcer conditions complicated by hemorrhage. Previous validation in anticoagulated populations suggests sensitivities of 95.7% and specificities of 97.2% for similar code sets [3].

- **Falls and Fractures:** Due to the high granularity of ICD-10 codes for orthopedic injuries, direct code matching is often insufficient. We adopted the hierarchical definitions from the Clinical Classifications Software Refined (CCSR v2025) [1]. To address potential discrepancies between granular billing codes (e.g., `M84750A`) and truncated EHR codes (e.g., `M8475`), we implemented a recursive search strategy. We identified root ICD codes (e.g., `S02`) where the entire branch pertains to the target condition and included all descendant codes (e.g., `S02.1`, `S02.101A`) in the concept set.

All ICD-based definitions were translated to SNOMED via OMOP relationships to query the standardized data.

#### S3. Data processing and label derivation for MIMIC-IV

##### Access and Privacy

We utilize MIMIC-IV, a de-identified database containing over 65,000 ICU admissions and 200,000 emergency department visits. Access was granted following the completion of credentialing and a data use agreement (DUA), ensuring strict adherence to privacy safeguards and non-re-identification protocols.

##### Label Generation

We leverage the Athena vocabulary service to expand our adverse event (AE) root codes. For every target category, we identify all descendant codes by normalizing the roots (removing periods and capitalizing) and performing a prefix match against the full set of concepts in the Athena `ICD10CM` and `ICD10GM` vocabularies.

A patient is assigned a positive label for category $j$ ($y_j = 1$) if their diagnosis list contains *any* code matching this expanded set of descendants. Patients with no matching codes for any category are labeled as *None* (or "no").

**Feature Extraction**

- **Medications:** Drug exposures are extracted from the Electronic Medicine Administration Record (EMAR). We normalize medication names (lowercasing and trimming whitespace) and aggregate them into a unique set per patient. These unique strings are mapped to integer tokens to form the `administered_drugs` sequence.
- **Complications:** We map source diagnoses (ICD-9 and ICD-10) to standard SNOMED Condition concepts using the "Maps to" and "Maps to value" relationships in Athena.

  *Exclusion Criteria:* To prevent data leakage, we strictly exclude any diagnosis code (and its descendants) that falls within the expanded AE definitions described above. Only non-AE diagnoses are mapped. The remaining unique SNOMED concept names are tokenized to form the `complications` sequence.
- **Cohorts:** We merge patient demographics (age, sex), drug sequences, complication sequences, and AE labels. We filter for data completeness, retaining only records that contain at least one valid medication entry and one valid complication entry.

#### S4. Generative AI Augmentation Details

**Conditional VAE (CVAE) Baseline.** The CVAE was trained exclusively on the subset of patients with positive AE labels ($\mathcal{D}_{pos}$). We represent each patient as a fixed-size vector $x \in \mathbb{R}^D$ constructed by concatenating normalized age, one-hot encoded sex, and multi-hot vectors representing drugs and complications. The AE labels are encoded as a condition vector $c \in \{0, 1\}^C$. The model consists of an encoder $q_\phi(z \mid x, c)$ and a decoder $p_\theta(x \mid z, c)$, where $z \in \mathbb{R}^{64}$ is the latent variable. The objective function is:

$$\mathcal{L} = \mathcal{L}_{\text{recon}}(x, \hat{x}) + \beta \cdot D_{KL}(q_\phi(z \mid x, c) \| \mathcal{N}(0, I))$$

The reconstruction term $\mathcal{L}_{\text{recon}}$ is a weighted sum of Binary Cross Entropy (for binary features) and Mean Squared Error (for age, weighted by $\lambda_{age} = 10$). We set $\beta = 1$ and train for 300 epochs using Adam ($\alpha = 10^{-3}$, batch size 256).

**Conditional GAN (CGAN) Baseline.** The CGAN consists of a generator $G(z, c)$ and a discriminator $D(x, c)$. The generator maps a noise vector $z \sim \mathcal{N}(0, I)$ and label condition $c$ to a synthetic feature vector $\hat{x}$. The discriminator attempts to distinguish between real pairs $(x, c)$ and synthetic pairs $(\hat{x}, c)$. Both networks are Multi-Layer Perceptrons (MLPs). We train for 5000 epochs using Adam ($\alpha = 10^{-4}$, $\beta_1 = 0.5, \beta_2 = 0.999$, batch size 1024) with a binary cross-entropy objective. During augmentation, we sample conditions $c$ from the empirical distribution of the training set.

### S5. Classical Classifier Baselines

#### S5.1 Data representation and Experimental Setup

We evaluate three classical machine learning models: decision tree, random forest, and XGBoost. Unlike the TASER-AE pipeline, we tune the classical baselines locally on each dataset to establish strong dataset-specific benchmarks.

- **MEARS Protocol:** We use the fixed stratified split (64/16/20). Hyperparameters are tuned via grid search on the validation split to maximize Macro F1. The optimal configuration is refit on the train+validation union and evaluated on the test set.
- **MIMIC-IV Protocol:** We split MIMIC-IV using the same ratios and stratification logic. We perform an independent grid search on the MIMIC validation split, refit on the union, and evaluate on the MIMIC test set.

#### S5.2 Hyperparameter Grids

##### Decision Tree

- `criterion` ∈ {`gini`, `entropy`}
- `max_depth` ∈ {`None`, 10, 50}
- `min_samples_split` ∈ {2, 5, 10}
- `min_samples_leaf` ∈ {1, 2, 4}
- `max_features` ∈ {`None`, `sqrt`, `log2`}

##### Random Forest

- `n_estimators` ∈ {100, 200, 500}
- `max_depth` ∈ {`None`, 10, 50}
- `min_samples_split` ∈ {2, 5, 10}
- `min_samples_leaf` ∈ {1, 2, 4}

**XGBoost** We rely on the default architectural parameters (depth, subsampling, regularization) but tune the number of boosting rounds. We employ early stopping with a patience of 10 rounds, monitoring Macro F1 on the validation split to determine the optimal iteration count.

#### S6. Supplementary Figures

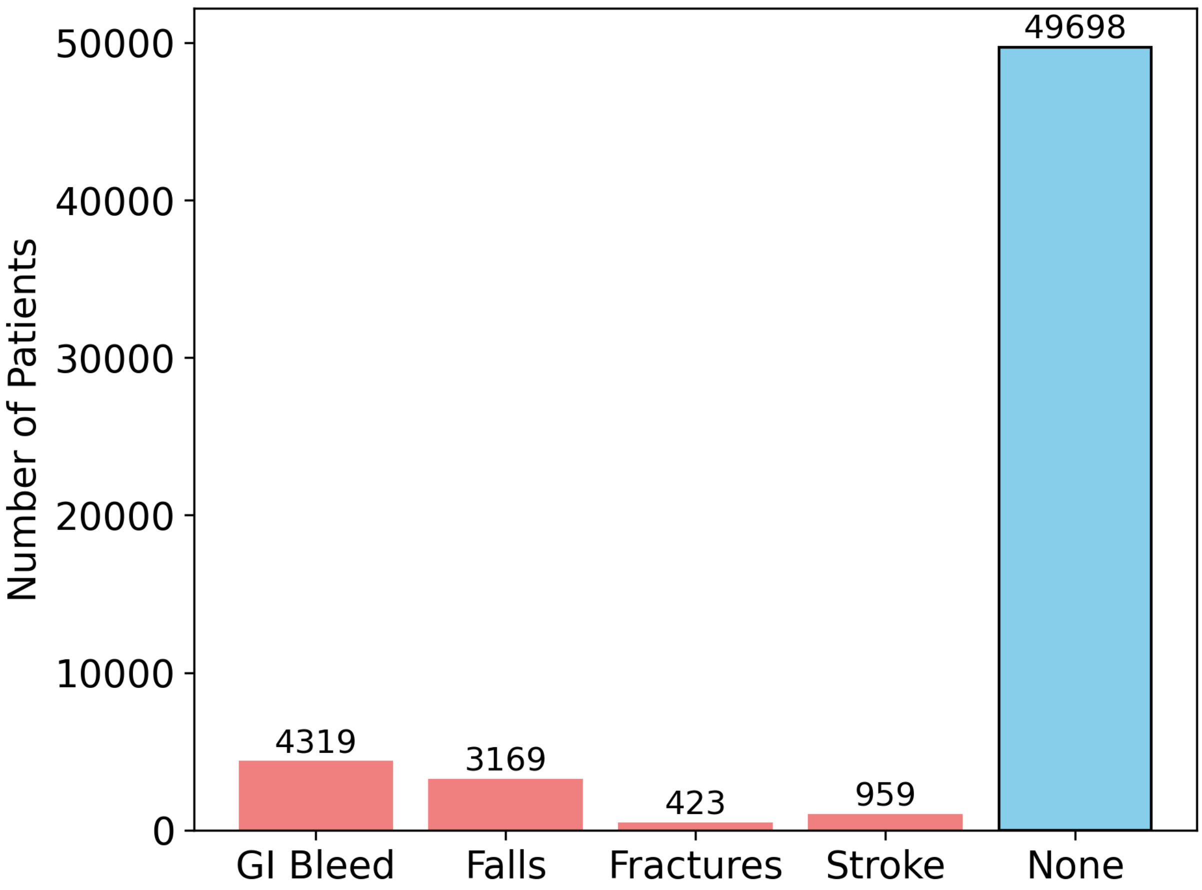


Figure S1: Patient counts for each adverse event label in MEARS.

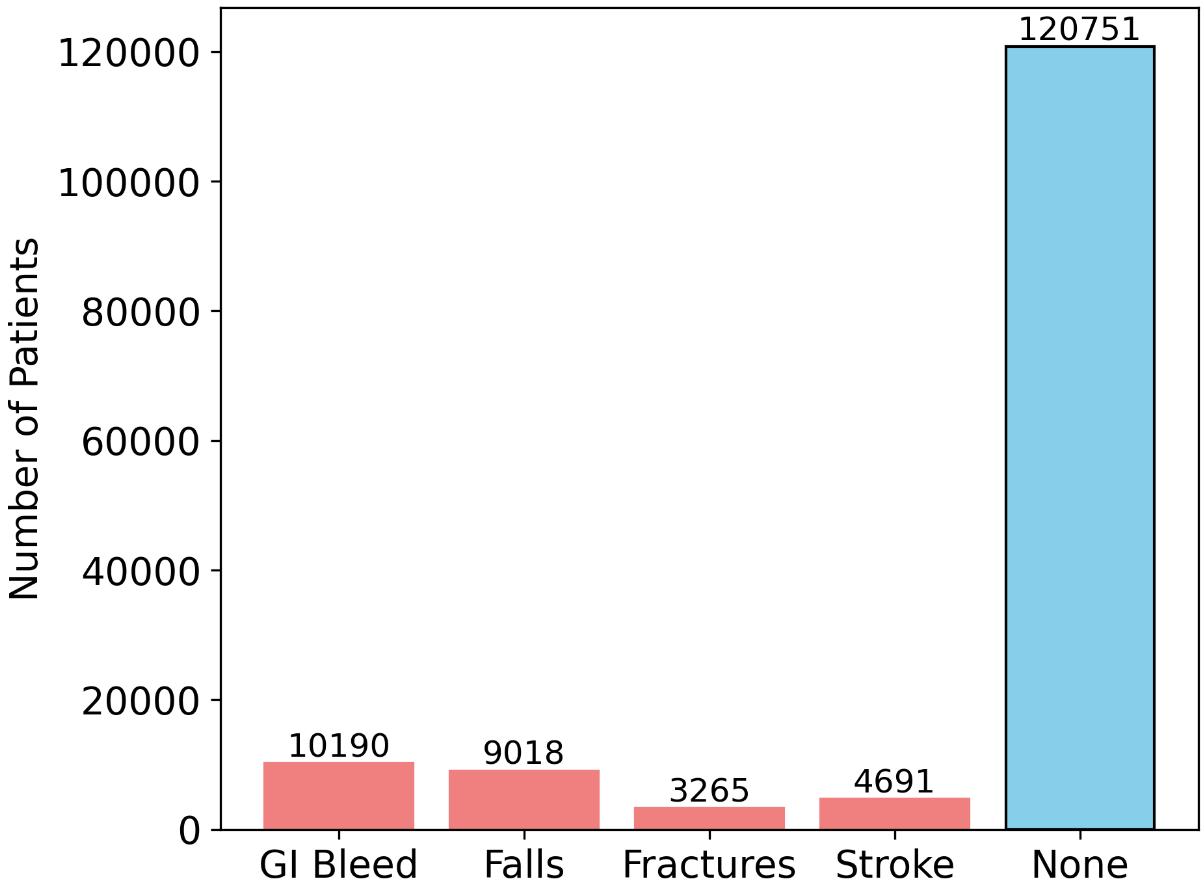


Figure S2: Patient counts for each adverse event label in MIMIC-IV.

#### S7. Supplementary Tables

Table S1: Performance of Classical Models on MIMIC-IV Minority Classes

| **Falls and Fractures** | | | | | | |
|---|---|---|---|---|---|---|
| **Model** | **Falls** | | | **Fractures** | | |
| | Precision | Recall | F1 | Precision | Recall | F1 |
| Decision Tree | 0.0890 | **0.0996** | **0.0940** | 0.0721 | **0.0838** | **0.0775** |
| Random Forest | 0.0000 | 0.0000 | 0.0000 | **1.0000** | 0.0029 | 0.0058 |
| XGBoost | **0.1667** | 0.0007 | 0.0015 | 0.2889 | 0.0376 | 0.0665 |

| **GI Bleed and Stroke** | | | | | | |
|---|---|---|---|---|---|---|
| **Model** | **GI Bleed** | | | **Stroke** | | |
| | Precision | Recall | F1 | Precision | Recall | F1 |
| Decision Tree | 0.1357 | **0.1547** | **0.1446** | **0.0694** | **0.0825** | **0.0754** |
| Random Forest | 0.4315 | 0.0361 | 0.0666 | 0.0000 | 0.0000 | 0.0000 |
| XGBoost | **0.4500** | 0.0258 | 0.0488 | 0.0000 | 0.0000 | 0.0000 |

| **Averages** | | | | | | |
|---|---|---|---|---|---|---|
| **Model** | **Macro Average** | | | **Minority Average** | | |
| | Precision | Recall | F1 | Precision | Recall | F1 |
| Decision Tree | 0.2470 | **0.2535** | **0.2499** | 0.0915 | **0.1051** | **0.0979** |
| Random Forest | **0.4571** | 0.2073 | 0.1985 | **0.3579** | 0.0097 | 0.0181 |
| XGBoost | 0.3519 | 0.2122 | 0.2073 | 0.2264 | 0.0160 | 0.0292 |

### References

[1] Agency for Healthcare Research and Quality (AHRQ). Clinical Classifications Software Refined (CCSR). https://hcup-us.ahrq.gov/toolssoftware/ccsr/ccs_refined.jsp, Dec. 2025. Accessed 2 December 2025.

[2] J. A. Columbo, N. Daya, L. D. Colantonio, Z. Wang, K. Foti, H. I. Hyacinth, M. C. Johansen, R. Gottesman, P. P. Goodney, V. J. Howard, P. Muntner, A. L. C. Schneider, E. Selvin, and C. W. Hicks. Derivation and Validation of ICD-10 Codes for Identifying Incident Stroke. *JAMA Neurol.*, 81(8):875–881, Aug. 2024. ISSN 2168-6149. doi: 10.1001/jamaneurol.2024.2044.

[3] C. Joos, K. Lawrence, A. E. Jones, S. A. Johnson, and D. M. Witt. Accuracy of ICD-10 codes for identifying hospitalizations for acute anticoagulation therapy-related bleeding events. *Thromb. Res.*, 181: 71–76, Sept. 2019. ISSN 0049-3848. doi: 10.1016/j.thromres.2019.07.021.

[4] Observational Health Data Sciences and Informatics (OHDSI). ATLAS. https://github.com/OHDSI/Atlas, Dec. 2025. Accessed 2 December 2025.

[5] Observational Health Data Sciences and Informatics (OHDSI). OMOP Common Data Model. https://ohdsi.github.io/CommonDataModel, Nov. 2025. Accessed 2 December 2025.

[6] Pennsylvania Department of Aging. Pharmaceutical Assistance – PACE Program. https://www.pa.gov/agencies/aging/aging-programs-and-services/pace-program, Dec. 2025. Accessed 2 December 2025.

[7] U.S. Department of Veterans Affairs. CIPHER. https://phenomics.va.ornl.gov/web, Dec. 2025. Accessed 2 December 2025.
